# Glucagon-Like Peptide 1 Receptor Agonists and Cardiovascular Disease Risk: Findings from Real-World Data using AI-Powered Outcomes

**DOI:** 10.1101/2024.09.17.24313797

**Authors:** Shivaani Prakash, Jeffrey Coleman, Jakob Steinfeldt

**Affiliations:** Dandelion Health; Independent contractor; Pheiron

## Abstract

**Background:** The SELECT trial showed cardiovascular benefits of glucagon-like peptide-1 receptor agonists (GLP-1 RAs) in obese patients with cardiovascular disease; however, real-world data (RWD) on this benefit remain limited. This study used an artificial intelligence (AI)-generated algorithm and multimodal RWD to evaluate the impact of GLP-1 RAs on cardiovascular disease risk in a population of obese patients with and without preexisting cardiovascular disease.

**Methods:** Using data from the Dandelion Health RWD library, an Emulated SELECT Cohort was created to include obese patients similar to those in the SELECT trial, but with and without preexisting cardiovascular disease. An AI algorithm developed by Pheiron that used 12-lead electrocardiograms (ECGs) as a predictive biomarker for the risk of major adverse cardiovascular events (MACE) was validated and used to derive MACE risk scores for the Emulated SELECT Cohort. These outcomes were compared over time between patients who used GLP-1 RAs and non-users using inverse-probability weighted linear regression models, adjusting for key covariates.

**Results:** Out-of-sample validation showed high predictive accuracy of the AI algorithm, with ROC AUCs of 0.81 for myocardial infarction (MI) and 0.75 for stroke. Increased risk scores from the algorithm were correlated with higher MACE incidence in RWD. In the Emulated SELECT Cohort of 20,795 patients, GLP-1 RA use was associated with significant attenuation of MACE risk, with reductions observed in percentile risk score for MI (4%; p<0.001) and stroke (3.6%; p<0.001) per year of use. Differences in GLP-1 RA and non-GLP-1 users were evident as early as 1.7 years, with a 15-20% difference in absolute MACE risk scores between GLP-1 RA users and non-users observed by the end of the study.

**Conclusion:** An AI algorithm using 12-lead ECGs accurately predicted MACE risk and could be used to model risk attenuation associated with GLP-1 RA use. Using this outcome, we find that GLP-1 RA use was associated with significant reductions in MI and stroke risk, in a broader population and within a shorter timeframe than the SELECT trial. These findings suggest potentially significant cardioprotective benefits of GLP-1 RAs in real-world settings and demonstrate a proof-of-concept for utilizing clinical AI to understand these benefits.

## INTRODUCTION

The uptake of glucagon-like peptide-1 (GLP-1) receptor agonists (GLP-1 RAs) has increased dramatically in the last four years, particularly among patients with type 2 diabetes (T2D) and/or obesity [Li 2024], as these agents have proven effective in promoting glycemic control and weight loss. [Sattar 2021] GLP-1 RAs mimic the activity of the incretin hormone GLP-1, which in turn stimulates postprandial insulin secretion and inhibits glucagon release, thereby improving postprandial glycemic control. [Anderson 2018; Holst 2007; Sandoval 2015] Subsequently, it was observed that agonism at the GLP-1 receptor, along with a delay in gastric emptying, reduced food intake and body weight. [Semaglutide PI 2024] This led to several GLP-1 RAs receiving US Food & Drug Administration (FDA) approval for the reduction of excess body weight and maintenance of weight reduction, including among adult patients with weight-related comorbidities.

Given the high cardiovascular risk in patients with obesity and diabetes, the FDA has also issued guidance mandating cardiovascular outcomes trials for GLP-1 RAs and other antiglycemic drugs for patients at increased risk of adverse cardiovascular outcomes. [McGuire 2019] Large randomized controlled trials such as the LEADER trial of liraglutide and the SUSTAIN-6 trial of semaglutide demonstrated that administering GLP-1 RAs to patients with diabetes and severe pre-existing cardiovascular disease significantly reduces the risk of major adverse cardiovascular events (MACE) including cardiovascular death, heart attacks, and strokes. [Marso 2016; Marso 2016b] As a logical next step, the SELECT (Semaglutide Effects on Cardiovascular Outcomes in People with Overweight or Obesity) trial examined the effects of a GLP-1 RA on overweight or obese patients with severe pre-existing cardiovascular disease, but without a history of diabetes.[Lincoff 2023] As in patients with diabetes, SELECT demonstrated that administering GLP-1 RAs to these patients reduced their risk of experiencing a MACE by 20% (p<0.05) as compared to a placebo group, after a mean follow-up time of 39.8 months. In March 2024, the FDA approved use of semaglutide to reduce MACE among adult patients with established cardiovascular disease who are either overweight or obese. [FDA Website 2024]

Although randomized controlled trials (RCTs) such as SELECT have long been considered the gold standard for isolating clinical effects with reduced bias, there remains limited real-world evidence on the potential cardioprotective benefits of GLP-1 medications in the patients who are actually being treated. This is partly because MACE are relatively rare, occurring in about 3% of the adult U.S. population annually. [Tsao 2023] In order to study occurrences of MACE, larger cohorts and multiple years of follow-up are required to detect treatment effects. Because both real-world research and clinical trials typically rely on tracking actual MACE occurrences, which can take years to observe, they often do not gauge the early appearance and progression of risk factors that might eventually lead to these events. [Kostis 2020] Subclinical changes that could be detected earlier, such as changes in MACE *risk*, are also difficult to assess using traditional patient health data such Electronic Health Records (EHR) and payer claims, as these are often designed for administrative purposes and thus may lack the clinical granularity to evaluate patient health comprehensively. These data sources may miss early indicators of health outcomes beyond outcomes explicitly coded in billed claims such as those for a MACE.

Assessing MACE risk in a more clinically meaningful way requires detailed, long-term data chronicling the incremental accumulation of risk factors that develop long before a MACE would occur. Moreover, in the case of SELECT, only patients with established cardiovascular disease were included. [Lincoff 2023] This strict inclusion criteria limited the generalizability of the SELECT findings to broader, real-world populations who are actually treated, or who might have benefited from a GLP-1 RA if it were available to them. Identifying subtle risk signals as they arise in a broader population would make early intervention to prevent MACE more feasible and cost-effective, by reducing both patient morbidity and the costs of treating acute cardiovascular disease events.

An AI-based approach to clinical research combined with multimodal real-world data (RWD) might help circumvent some of the obstacles posed by RCTs and current real-world evidence approaches. Multimodal RWD – raw unstructured data like physician notes, images, and waveforms, representing approximately 80% of data captured in the clinical setting – can provide deep insights into patient health. [Chen 2023/p3/col1/pars2&4; Kong 2019/p1/col1/par3] These rich data sources, however, have historically been inaccessible to researchers due to the time and cost required to interpret such data types at scale. [Kong 2019/p1/col1/par2; Rajkomar 2018/p1/col1/par2/p2/col1/pars4-5] Having cardiologists review tens of thousands of electrocardiograms (ECGs), for example, would be prohibitively expensive, and would produce narrower results than a well-trained algorithm could.

[Hernandez-Boussard 2019/p1190/col1/par2/p1191/col1/par2] AI models can analyze extremely large, diverse and unstructured datasets in a fraction of the time and cost required for RCTs. [Chen 2023/p3/col1/pars4-5; Hernandez-Boussard 2019/p1190/col1/par2/p1191/col1/par2; Rajkomar 2018/p1/col1/par2/p2/col1/pars4-5] This can help researchers avoid the recruitment challenges, adherence issues, and participant dropouts that can make it harder to obtain and maintain the type of sufficiently large study population necessary for meaningful statistical comparisons. [Chopra 2023]

By using RWD, AI can enhance the generalizability of findings, making them more applicable to broader patient populations. A well-designed AI-based study can evaluate treatment effects longitudinally throughout treatment to identify risk signals early when preventive care may still be possible. In other words, AI-based research can help examine the effects of GLP-1 RAs with the methodological rigor of an RCT, but earlier and in the population currently receiving these therapies in actual clinical practice rather than under controlled conditions at a study site.

In this manuscript, we present a proof-of-concept study using a multimodal patient RWD library (Dandelion Health, Syosset NY) to validate whether a novel AI-generated biomarker can accurately predict the risk of MACE and then utilized the validated algorithm for understanding the benefit of GLP-1 RAs. Using opportunistic 12-lead ECG waveforms, we investigated whether GLP-1 RA use impacts risk of MACE over time by comparing AI-predicted MACE risk scores between obese patients both with or without severe preexisting cardiovascular illness who used GLP-1 RAs and similar patients who did not use them. The proposed advantage of using this biomarker is that it identifies subtle changes in cardiovascular status that might increase cardiovascular risk but be apparent long before a MACE requires critical intervention instead of preventive care.

## METHODS

### Data Source

This study utilized a subset of Dandelion Health Data, a database of regularly updated patient-level multimodal clinical data (2016 to present) collected from a geographically diverse consortium of US non-academic healthcare systems. [Dandelion 2024] The database includes patient-level EHR data on demographics, diagnoses, procedures, health system encounters, medication orders and administration route, laboratory and diagnostic test orders and results (e.g., blood glucose and potassium) and vital signs (e.g., heart rate and blood pressure). These data are linked to unstructured data such as ECG waveforms, medical imaging (e.g., Computed Tomography scans, Magnetic Resonance Imaging, X-rays, Ultrasounds), associated radiology reports, echocardiograms and clinical notes, to create a longitudinal, multimodal dataset capturing the trajectory of clinical care.

Prior to extraction from a health care system, Dandelion data are de-identified via privacy-preserving methodologies that are specifically developed for each data type and approved by expert determination under the HIPAA Privacy Rule. [HHS 2022]

Once extracted, data are harmonized across healthcare systems into a unique data model through semantic processing and a data transformation layer, and then made available for analysis on the Dandelion platform. The data utilized for this study were primarily drawn from EHRs accessed in January 2024. The data included observations from January 2019 - October 2023 to ensure a contemporary dataset reflective of current clinical practices and recent GLP-1 RA uptake.

### Cohort Creation

Using the Dandelion data, we created an Emulated SELECT Cohort which was used to determine whether the cardioprotective benefits of GLP-1 RAs observed in the real-world setting are similar to those observed in the SELECT clinical trial population, but with modified inclusion criteria to assess a broader patient population.

Data for this cohort were first identified by applying the SELECT trial’s inclusion and exclusion criteria to the Dandelion dataset (Fig. 1). Eligible patients included those who had at least two outpatient encounters with a primary care physician or endocrinologist, at least 2 months apart, with the first encounter identified as the study entry. Patients included in the study were aged >45 years at study entry with evidence of obesity (based on body mass index [BMI] > 27 or prior diagnosis codes) but did not have a diagnosis of diabetes in their clinical records. Similar to SELECT, the Emulated Cohort excluded patients with high HgA1C values, end-stage kidney disease, acute pancreatitis, dialysis use and instances of myocardial infarction (MI), unstable angina, stroke or transient ischemic attack in the previous 60 days. However, unlike the SELECT trial population, the Emulated Cohort did not require patients to have severe pre-existing cardiovascular disease, defined as having had a previous myocardial infarction, stroke (ischemic or hemorrhagic), peripheral arterial revascularization procedure, or amputation due to atherosclerotic disease. With the inclusion of these patients, the Emulated SELECT Cohort allowed for evaluation of GLP-1 RAs in a broader, more diverse population of patients than was enrolled in SELECT.

**Figure 1.**
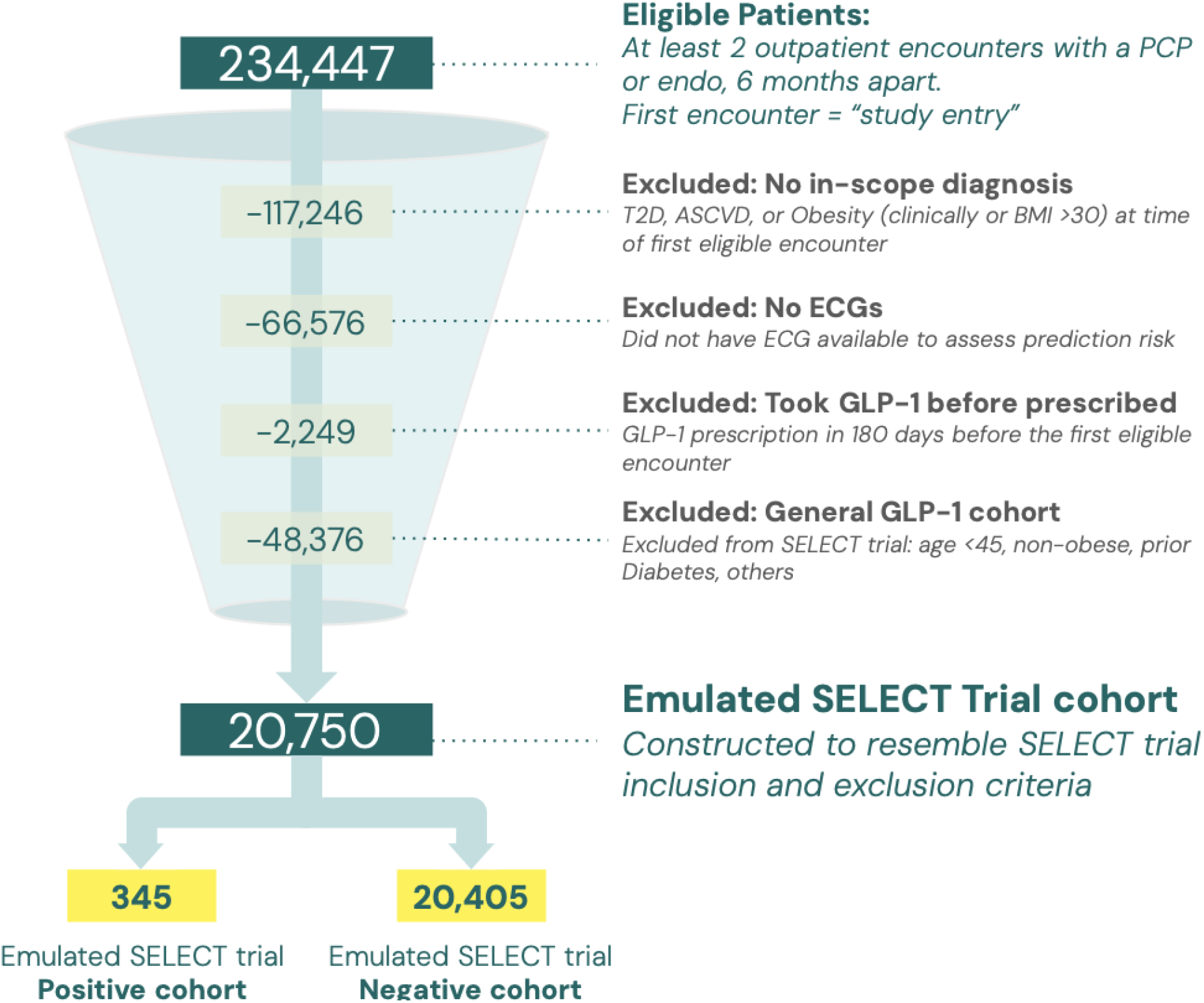
Overall Cohort Definition

Among eligible patients, we identified a Positive Group with at least one medication order for GLP-1 RAs following study entry and a Negative Group with no history of GLP-1 RA use in their clinical record. A dataset for these patients was constructed for analysis including diagnoses, medications, labs, vitals, procedures, demographics information, and all available ECGs in patient clinical records from January 2019 - October 2023.

The broadened inclusion criteria we utilized extended the timeframe across which treatment effects could be identified. In SELECT, using MACE as an outcome would not have been feasible if patients without cardiovascular disease had been enrolled because the incidence of MACE in this population would be low within the limited timeframe provided for an RCT. A very large sample size and long follow-up would have been required to detect significant treatment effects. The Emulated SELECT cohort, which also included patients without cardiovascular disease, essentially created an earlier baseline. Studying earlier treatment effects and using an AI algorithm that measures MACE risk allowed for assessment of patients who might experience cardioprotective benefits from GLP-1 RAs prior to developing more severe cardiovascular risk factors or experiencing a MACE that might not be detected within the time frame of a conventional clinical trial.

### Algorithm and AI-Generated Outcome Selection

The AI-generated primary endpoint selected for our study was the percentile rank-transformed scores for risk of two types of incident MACE – myocardial infarction and stroke – as predicted using an algorithm that interprets patients’ opportunistic 12-lead ECGs. [Pheiron 2024] The AI-biomarker algorithm, developed by Pheiron, leverages a fine-tuned neural network specifically trained to identify patterns in ECG input to predict cardiovascular outcomes with high sensitivity and specificity. Neural networks are designed to mimic the data processing activity of the brain, harnessing powerful hierarchical computing methods for pattern recognition and classification tasks. [Kumar 2022]

This algorithm is part of a platform that utilizes a proprietary AI-driven system to create phenotypic profiles specifically designed to predict clinical outcomes under various conditions and support drug development. [Pheiron 2024] The neural network within the platform, trained and fine-tuned on several hundred thousand ECGs, can mirror the clinician’s role in interpreting ECG data during an office visit. Because the algorithm analyzes ECG waveforms as the sole input, it can capture subclinical cardiovascular disease risk manifesting as irregular heart rhythms. This is possible even in patients who have not experienced MACE, had diagnostic test results indicating cardiovascular risk factors for such events (e.g., atherosclerosis, elevated blood pressure, high cholesterol), or shown visible symptoms of heart disease. This allows for evaluation of treatment effects occurring long before the normal baseline of a traditional clinical trial.

### Analysis: Algorithm Validation

Before utilizing the algorithm to predict MACE risk in our Emulated SELECT cohort, we wanted to confirm the accuracy of the algorithm by conducting our own out-of-sample validation. Using the Dandelion multimodal RWD dataset, we randomly selected 100,000 12-lead ECGs from adults aged ≥18 years who were *not* included in the Emulated SELECT cohort. ECGs with sampling rate below 250 Hz and those of poor quality (e.g., with artifacts or baseline wander) that the algorithm could not read were excluded from the final sample of 100,000. For patients whose ECGs were included in the validation dataset, we looked for any diagnosis of myocardial infarction or stroke occurring after ECG date in order to create binary flags for the two real-world ground-truth outcomes. Because both ECGs and diagnosis codes are available and longitudinally linked within the Dandelion datasets, leveraging them to perform this independent validation was straightforward and provided additional confidence in the value of the algorithm for our study.

We then ran the algorithm on our 100,000 validation ECGs and subsequently conducted two validation analyses for both myocardial infarction and stroke to assess algorithm accuracy and performance using our ground-truth outcomes. To achieve this, we calculated the Area Under Receiver Operating Characteristics Curve (ROC AUC), both overall and stratified by sex, age band, race/ethnicity, rurality (rural vs. urban), and median income decile (based on US Census Data for patients’ Census Tract) to ensure consistent performance and minimize bias. We also plotted the ground-truth outcome probability against percentile risk scores generated by the algorithm from ECG readings in order to assess concordance with real-world incidence of outcomes.

### Analysis: Evaluation of GLP-1 RA Impact

After validation, the algorithm was utilized to generate risk scores for all ECGs available for patients in the Positive and Negative groups of the Emulated SELECT trial cohort. We then used inverse probability-weighted (IPW) linear regression models to compare the aggregated predicted-risk scores for GLP-1 RA users versus non-users over time following study entry, adjusting for key variables such as demographics, comorbidities, medical procedures, medications, glucose levels, HbA1c, and BMI at baseline (study entry). The primary modeling specification included an interaction term between GLP-1 RA use and duration of GLP-1 RA use, to assess how the effectiveness of GLP-1 RAs might change over time. These analyses were conducted separately for the myocardial infarction and stroke risk score outcomes. Sensitivity analyses included testing propensity score-matched models, as well as covariate-adjusted difference-in-difference models assessing changes in MACE risk before and after GLP-1 RA initiation in the Positive and Negative groups for just those patients with serial ECG data pre- and post- GLP-1 RA initiation available.

Additional sensitivity analyses tested unadjusted models and covariate-only adjusted models, without IPW estimation applied.

## RESULTS

### Findings: Validation

Out-of-sample validation results successfully produced risk scores for all 100,000 ECGs in the validation sample. In the validation sample, 1.9% of respondents had a myocardial infarction following ECG and 1.8% of respondents had a stroke. We found a model ROC AUC of 0.81 for myocardial infarction and 0.75 for stroke, suggesting high accuracy of classification against ground truth outcomes as well as good generalizability and lack of overfitting to the specific validation dataset. In addition, ROC AUC results were consistent by age, race/ethnicity, gender, rurality and median income decile for both endpoints, with all ROC AUC values for subgroups within 5-10% of the overall model, suggesting no significant biases in algorithm performance.

The ground-truth outcome probability by risk score bins (Fig. 2) demonstrated concordance with real-world events; as the algorithm risk score percentile increased, so did the prevalence of actual MACE occurrences. The group of patients that the algorithm predicted to be highest at-risk for myocardial infarction (Fig. 2a) and stroke (Fig. 2b) were those experiencing MACE most frequently in the real-world. The rate was also non-linear (e.g. the slope increased at higher risk score percentiles), meaning that the more the algorithm *predicted* high risk of MACE, the more likely it was that MACE was actually *occurring* in the validation sample population.

**Figure 2:**
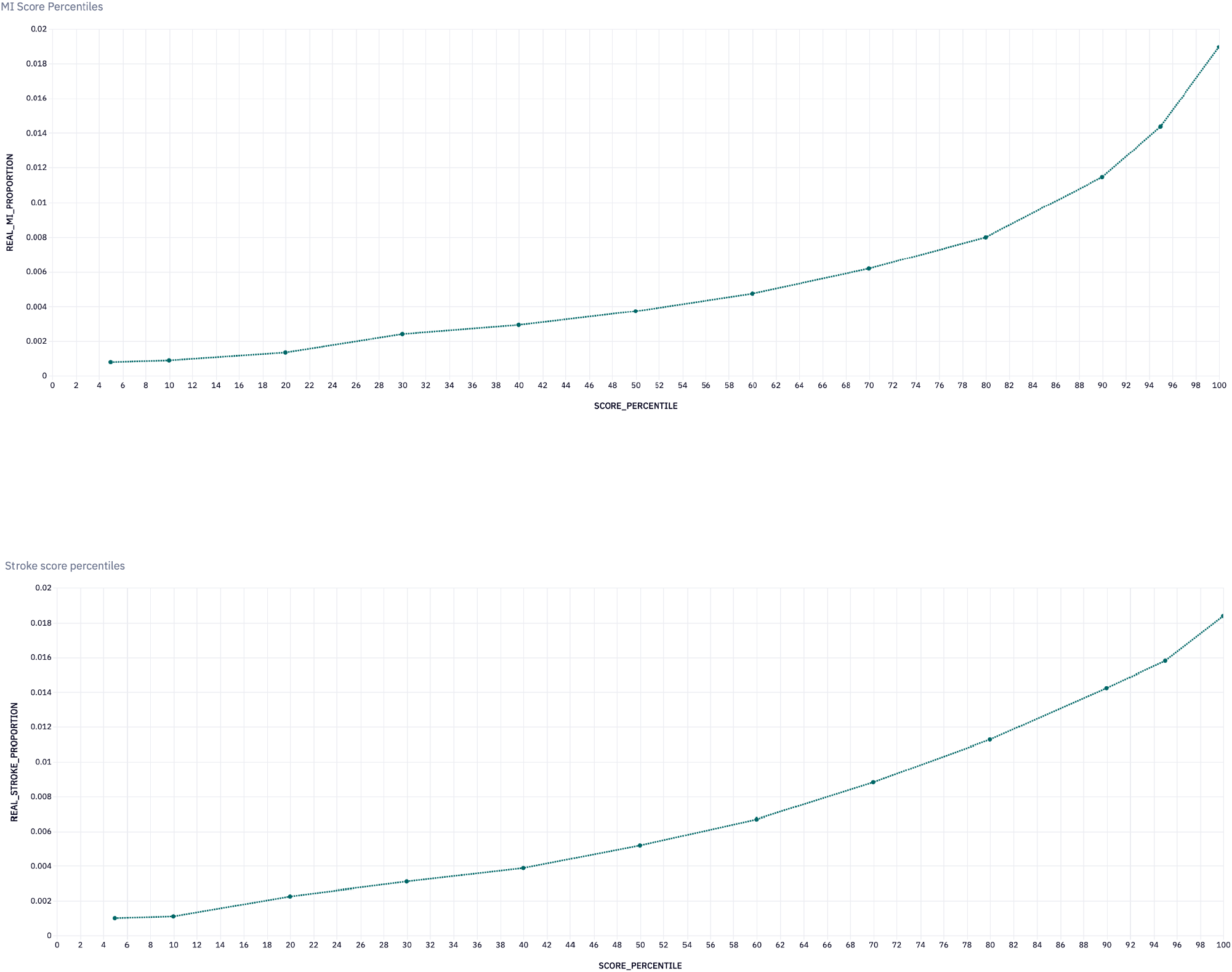
Validation Results from Out-of-Sample Validation **a:** Validation of AI-Derived Myocardial Infarction Risk Score Percentile by Incidence of M **b:** Validation of AI-Derived Stroke Risk Score Percentile by Incidence of Stroke

### Findings: Evaluation of GLP-1 RA Impact

#### Emulated SELECT Cohort Descriptive Results

Once modified inclusion/exclusion criteria were applied, the Emulated SELECT cohort included a total of 20,795 patients who had at least one 12-lead ECG waveform reading (Table 1). As in the SELECT trial, patients aged >45 years of age and patients with a BMI >27 and/or a diagnosis of obesity were included, while those with diabetes or recent cardiac events and severe kidney disease were excluded. The final Emulated SELECT cohort consisted of these 345 patients in the Positive Group and 20,450 patients In the Negative Group.

**Table 1.** Descriptive Characteristics of Emulated SELECT Trial Cohort.

| Characteristics | Emulated SELECT Trial<br>Positive Cohort<br>(n = 345) | Emulated SELECT Trial<br>Negative Cohort<br>(n = 20,405) |
| --- | --- | --- |
| **Age band, n (%)** |  |  |
| Aged 18-44 | N/A | N/A |
| Aged 45-64 | 286 (82.9) | 11423 (56.0) |
| Aged 65-80 | 58 (16.8) | 7415 (36.3) |
| Aged 80+ | 1 (0.3) | 1567 (7.7) |
| **Female, n (%)** | 261 (75.7) | 11234 (55.1) |
| **Race/ethnicity, n (%)** |  |  |
| White, Non Hispanic | 191 (55.4) | 11415 (55.9) |
| Hispanic | 65 (18.8) | 4329 (21.2) |
| Asian | 8 (2.3) | 1083 (5.3) |
| Black or African American | 25 (7.2) | 861 (4.2) |
| American Indian, Alaska Native, Native Hawaiian or Other Pacific Islander | 7 (2.0) | 206 (1.0) |
| Other or Multiple Race | 26 (7.5) | 1129 (5.5) |
| Unknown/Declined | 23 (6.7) | 1382 (6.8) |
| **BMI Range, n (%)** |  |  |
| BMI < 25 | 0 (0.0) | 21 (0.1) |
| BMI 25-29 | 5 (1.4) | 3473 (17.0) |
| BMI 30-39 | 153 (44.3) | 14175 (69.5) |
| BMI 40+ | 187 (54.2) | 2736 (13.4) |

There were some baseline differences evident in the Positive and Negative Groups in terms of composition. From our RWD, those in the Negative group tended to be older – 44% were aged 65 years and above, as compared to only 17% of the Positive group – and were more likely to be male. There were similar proportions of White Non-Hispanic and Hispanic patients in both groups, while the Positive group had a slightly higher proportion of Black patients than the Negative group (7.2% vs. 4.2%). Unsurprisingly, patients in the Positive cohort had a higher average BMI than those in the Negative group at baseline, with over half of Positive group patients having a BMI ≥40 at baseline, as compared to only 13% of the Negative group. These differences suggest that adjusting for baseline covariates, as well as testing a propensity-score matching empirical specification, were necessary in our modeling approach.

#### Modeling Results

We found that GLP-1 RA use was significantly associated with a reduced risk of both myocardial infarction and stroke in an IPW linear model that adjusted for observable covariates such as age, race/ethnicity, sex, BMI, and pre-existing comorbidities at baseline (Table 2). Notably, the magnitude of this risk reduction increased with duration of GLP-1 RA use, such that each additional year of GLP-1 RA use was associated with incremental risk reduction. The primary analysis, which compared percentile rank scores from the AI-derived outcomes, found that each additional year of GLP-1 use was associated with a 4% reduction in the risk of myocardial infarction (p<0.001) and a 3.6% reduction in the risk of stroke (p<0.001). Findings from the modeling for covariate-adjusted propensity score-matched subsets of the Positive and Negative cohorts were directionally similar to the main modeling specification, although the coefficient findings from the propensity score models did not reach thresholds for statistical significance, likely due to sample size limitations in propensity score matching. Encouragingly, results were directionally similar and strongly statistically significant for the covariate-adjusted difference-in-difference modeling, demonstrating that there is significant reduction in MACE risk over time across serial ECGs among Positive group patients as compared to Negative group patients.

**Table 2:**
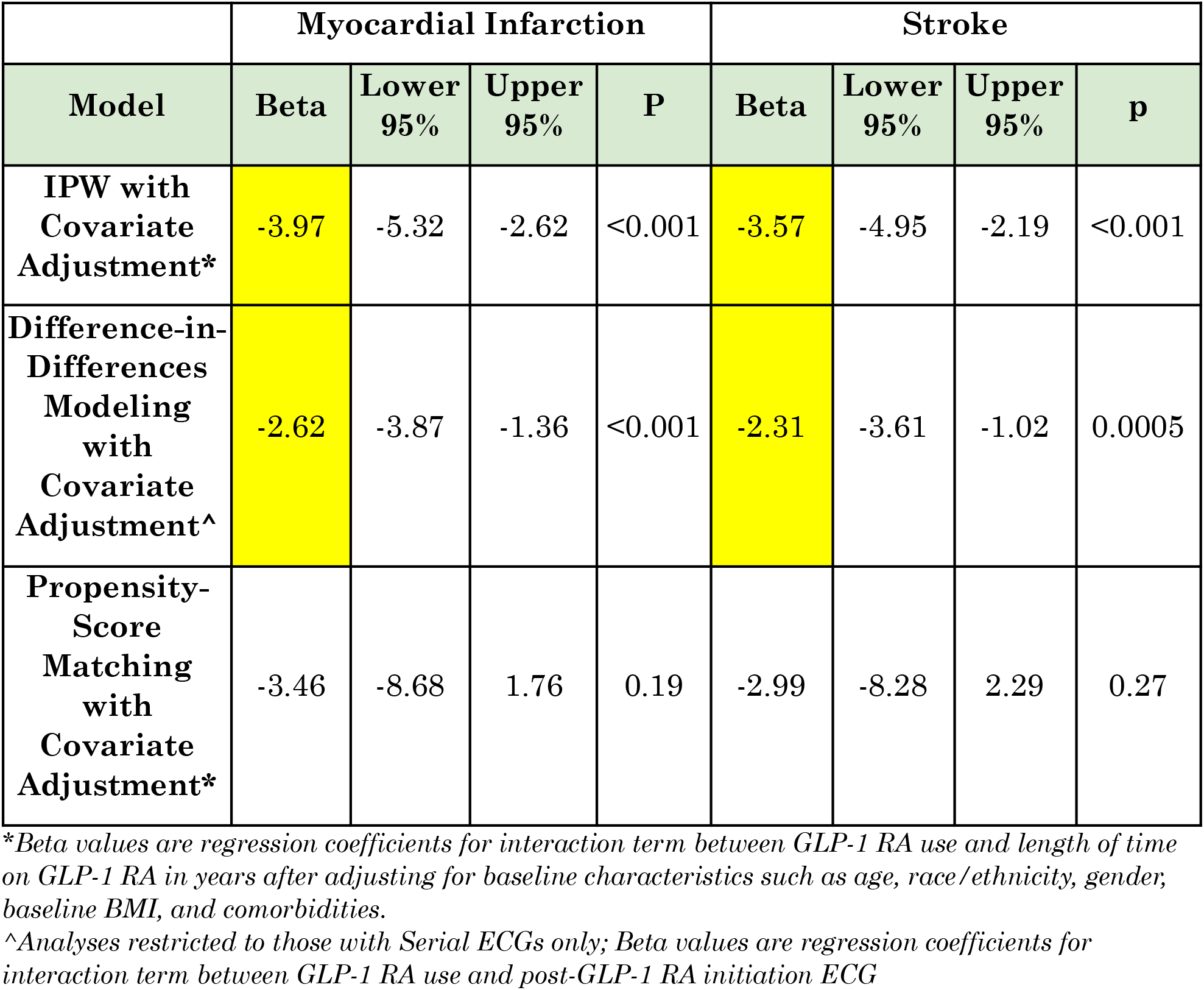
Modeling Results for Percentile Reduction in Risk of Myocardial Infarction and Stroke Over Time for GLP-1 Patients Compared to non-GLP-1 Patients Over Time.

|  | Myocardial Infarction |  |  |  | Stroke |  |  |  |
| --- | --- | --- | --- | --- | --- | --- | --- | --- |
| Model | Beta | Lower 95% | Upper 95% | P | Beta | Lower 95% | Upper 95% | p |
| IPW with Covariate Adjustment* | -3.97 | -5.32 | -2.62 | <0.001 | -3.57 | -4.95 | -2.19 | <0.001 |
| Difference-in-Differences Modeling with Covariate Adjustment^ | -2.62 | -3.87 | -1.36 | <0.001 | -2.31 | -3.61 | -1.02 | 0.0005 |
| Propensity-Score Matching with Covariate Adjustment* | -3.46 | -8.68 | 1.76 | 0.19 | -2.99 | -8.28 | 2.29 | 0.27 |

\*Beta values are regression coefficients for interaction term between GLP-1 RA use and length of time on GLP-1 RA in years after adjusting for baseline characteristics such as age, race/ethnicity, gender, baseline BMI, and comorbidities.
^Analyses restricted to those with Serial ECGs only; Beta values are regression coefficients for interaction term between GLP-1 RA use and post-GLP-1 RA initiation ECG

Results from unadjusted analyses and covariate-adjusted modeling without IPW estimation (*data not shown; available upon request*) were also directionally similar to the main modeling specifications but coefficients were not statistically significant.

While subclinical cardiovascular risk progression, as detected from the 12-lead ECGs, was evident in both the Positive and Negative groups over time, the rate of progression was notably slower in the Positive group. The attenuation of risk of myocardial infarction (Figure 2) and stroke (Figure 3) was statistically significant between GLP-1 RA users and non-users as early as 1.7 years after initiating GLP-1 RA therapy. This timeline was likely earlier than when significant differences in MACE incidence would typically be observed, whether in real-world settings or within a clinical trial context. At 36 months after GLP-1 RA initiation, a 15-20% difference in absolute MACE risk scores was observed between the Positive and Negative groups. This roughly corresponds with the approximately 20% cumulative reduction in MACE incidence with GLP-1 RA patients as compared with the placebo group over the 39-month follow-up period reported in the SELECT trial.

**Figure 3:**
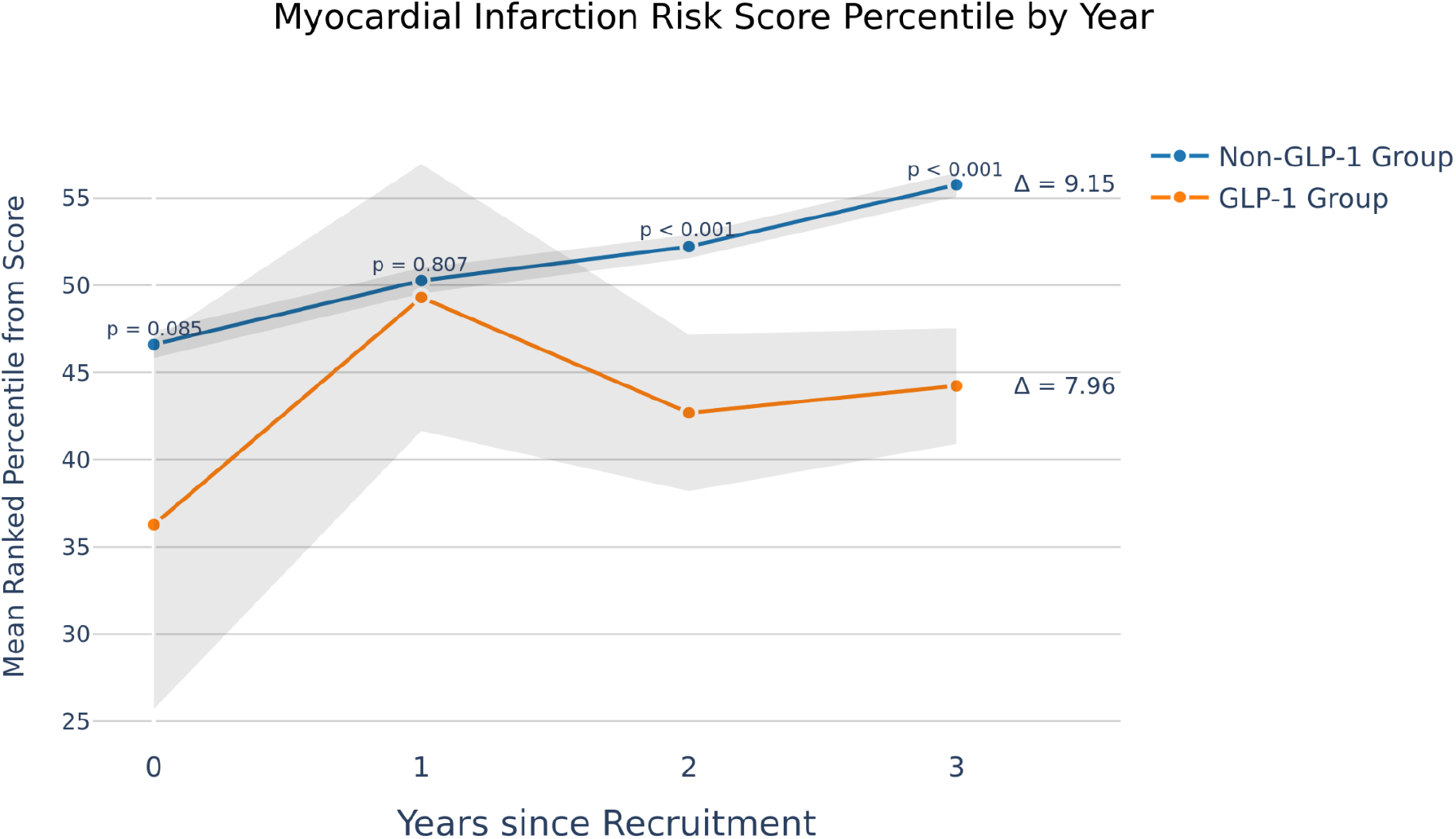
ECG-derived Myocardial Infarction Risk Score Percentile for GLP-1 Group and Non-GLP-1 Group in Emulated SELECT Cohort over time

**Figure 4:**
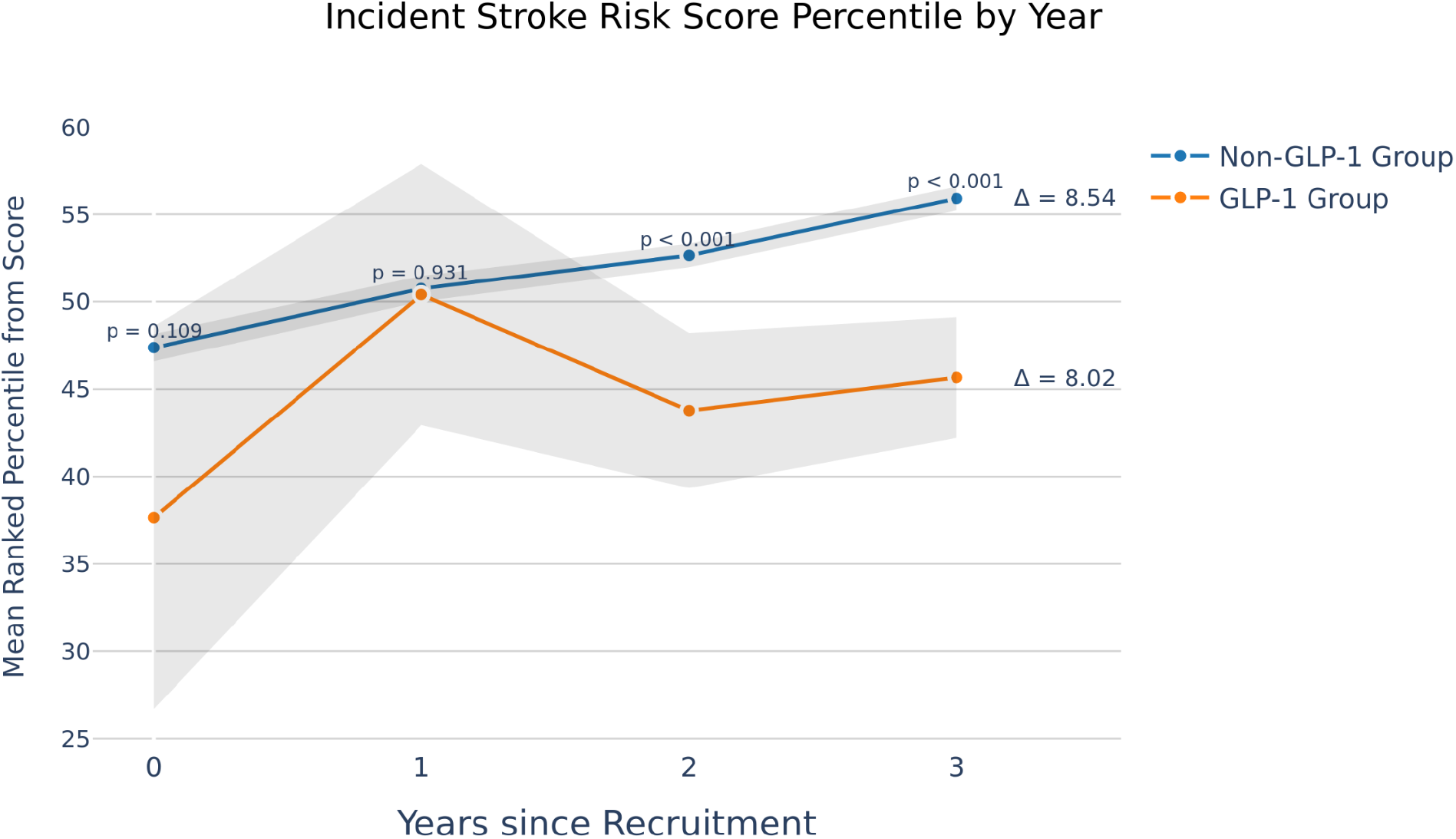
ECG-derived Incident Stroke Risk Score Percentile for GLP-1 Group and Non-GLP-1 Group in Emulated SELECT Cohort over time

## DISCUSSION

### Overview of Findings

We conducted a proof-of-concept study with the objective of validating an AI-derived endpoint and leveraging it to assess the potential benefit of a therapeutic class of medications using RWD from a longitudinal multimodal clinical dataset.

Opportunistic 12-lead ECGs were utilized as an input to an algorithm to derive an AI-generated predictive biomarker that quantified MACE risk over time. First, we used out-of-sample validation to demonstrate that the model could accurately assess MACE risk as compared to ground-truth outcomes. We then assessed the real-world cardioprotective benefit of GLP-1 RAs over time by comparing the AI-predicted MACE risk scores from ECGs of patients taking GLP-1 RAs to patients not taking a GLP-1 RAs, in a population that emulated the SELECT trial but did not exclude patients with preexisting cardiovascular disease. Our purpose in studying this population was to determine the extent to which findings from a large clinical trial conducted using standard methods could be applicable more broadly based on real-world treatment patterns.

In our study, the use of an AI-powered model offered significant advantages over a traditional clinical trial by expanding the scope of treatment evaluation to a broader population of potential patients who could be evaluated more readily. Traditional clinical trials like SELECT typically focus on MACE as the primary endpoint, [Marso 2016; Marso 2016b; Lincoff 2023] which inherently limits the inclusion of those with less prominent risk factors who are also less likely to experience a MACE within the typical time frame of a clinical trial. This approach makes it challenging to observe meaningful results in the absence of an acute event. As a result, these populations are often excluded from trials, leading to a narrower understanding of a treatment’s potential benefits.

In contrast, our AI-driven approach leverages multimodal RWD and advanced predictive models to provide an outcome measure that predicts risk of future MACE rather than relying on the actual occurrence of critical clinical events. Unlike MACE occurrence, which signals an adverse and often late-stage cardiovascular event, ECGs are regularly used diagnostic tools that generate findings that are not intrinsically negative. They can detect subtle changes in subclinical cardiovascular risk over time, even in patients who are at low or immediate risk of a MACE or in earlier stages of cardiovascular disease progression. [Fu 2024; Lih 2020] When combined with a multimodal dataset, this allows for the assessment of GLP-1 RAs in a broader, more diverse population, providing insights into how these treatments might prevent the progression of cardiovascular disease long before severe outcomes occur. Prioritizing a subclinical disease marker may help shift patient care away from secondary prevention to earlier, more proactive primary prevention, in a population that could greatly benefit from it. The significant efficacy that was demonstrated in the Emulated SELECT cohort suggests that clinical trials may underestimate the population that may potentially benefit from GLP-1 RAs.

Consistent with this hypothesis, our study demonstrated that GLP-1 RAs effectively slowed the progression of subclinical cardiovascular risk, with statistically significant reductions in myocardial infarction and stroke risk becoming evident as early as 1.7 years after treatment initiation. This finding suggests that meaningful differences in MACE risk may become evident sooner than typically observed in either real-world settings or clinical trials, and that starting therapy early, before cardiovascular disease is manifest, may afford important MACE risk reduction compared with waiting until risk factors become severe or cardiovascular disease manifests. Our findings indicate that GLP-1 RAs may be effective as primary prevention of cardiovascular disease, compared to the SELECT trial, which primarily evaluated GLP-1 RAs for secondary prevention.

Encouragingly, the findings in our Emulated SELECT cohort align with the original SELECT trial, where a 20% reduction in MACE risk was reported over 39 months. In our real-world analysis, we observed similar reductions in MACE risk scores for GLP-1 RA users over 36 months. While outcomes in the Emulated SELECT cohort and the SELECT trial are not directly comparable, the similarity suggests that the findings from our predictive model, based on RWD, are directionally consistent with those observed in the SELECT Trial. Beyond this, differences in cohort size and makeup highlight the extent to which clinical trial data may dramatically underestimate the therapeutic potential of GLP-1 RAs. Future research should clarify how differences between real-world and clinical trial treatment patterns might affect efficacy.

### Proof-of-Concept: Value of AI-Derived Outcomes

MACEs are relatively rare, typically occurring in 3% of the general US adult population in any given year. [Suying 2019] However, the risk of experiencing a recurrent MACE after having had a previous event is much higher than in the general population. Cardiovascular clinical trials that use MACE as an endpoint therefore tend to recruit these patients who have had a previous event, and require large cohorts observed over multiple years of follow- up in order to achieve sufficient statistical power to distinguish effects with active treatment from those in a control group. While primary prevention of MACE could benefit a greater population of patients, expanding the SELECT trial to this population would likely have required a population roughly 2.5-3 times larger than the one recruited, with a substantial corresponding increase in costs.

Given these logistical concerns, the dual benefit of not relying exclusively on MACE and using 12-lead ECGs as a predictive biomarker lies in the potential for earlier intervention in a range of patients not typically evaluated in clinical trials. While MACE measures only capture treatment effectiveness in trials after severe events have occurred, use of ECGs enables the identification of early warning signs, potentially allowing clinicians to intervene before adverse outcomes manifest. This shift from a reactive to a proactive approach would enhance patient care, potentially reducing the health and societal burdens of cardiovascular disease by preventing it rather than responding to it.

Moreover, utilizing 12-lead ECGs as a predictive marker for MACE risk not only allows for earlier detection in a larger population but also may eventually extend data collection outside of the clinic setting into everyday life. The advent of wearable 12-lead ECG technology represents a significant advancement in cardiovascular monitoring, consistent with the broader trend toward wearable devices designed to gauge other health parameters such as blood glucose levels and exercise levels [Duncker 2021/p11/par3-4; p17/par1]. Traditionally, output from 12-lead ECGs required interpretation by highly trained clinicians, which has limited their usefulness outside the clinic setting [Bouzid 2022/p2/par 1]. Wearable 12-lead ECG devices can overcome this limitation by bringing comprehensive heart monitoring directly into everyday life, offering continuous and real-time observation of cardiovascular function that may not be evident at all times, including during a clinic visit. The development of automated ECG-interpretation algorithms, such as the AI-enabled risk classification algorithm leveraged in our study, should improve the precision of ECG analysis [Bouzid 2022/p9/par3]. In short, an AI-driven approach to analyzing ECG data can allow for continuous risk assessment, earlier detection of issues that require redress, and round-the-clock monitoring [Dunker 2021/p17/par1], all of which could likely further contribute to cardioprotective therapeutic benefits.

Notably, the use of algorithms in a clinical context to predict or classify outcomes is not meaningful without the ability to validate the algorithm’s performance against ground-truth outcomes data. As of late, many clinical algorithms are trained on limited subsets of data that are drawn from a single modality, such as ECGs or CTs. The ability to link the findings from an algorithm directly to diagnosis codes, procedure codes, vitals and lab measurements in the same patient’s clinical record to validate the accuracy and degree of bias in an algorithm’s out-of-sample performance is crucial to have confidence in its performance for true clinical decision making. In this proof-of-concept study, we have demonstrated a process by which one can select and validate such an algorithm before using it for a more rigorous real-world evidence analysis.

### Strengths and Limitations

This study has numerous strengths to highlight. Our analysis included a Positive (GLP-1 RA treatment) and Negative (control) cohort that was followed longitudinally (median follow-up time available: 22 months), with a rich range of covariates from source-of-truth clinical data to select patients by implementing and removing different clinical trial inclusion and exclusion criteria. In clinical trials, selecting patients based on clinical criteria can be a tedious, manual exercise in reviewing clinical data, but the multimodal nature of the dataset in use allowed us to select the appropriate patients extremely quickly for a well-powered primary analysis. The multimodal aspect also allowed us to link ECGs to the real-world longitudinal clinical record for patients to both complete a separate validation exercise of the algorithm to generate the outcome and to conduct the Emulated SELECT trial analysis.

Our AI-derived outcome is based on an objective read of raw ECG waveforms and is less likely to suffer from the same degree of measurement error or bias as self-reported measures, or even some administratively-captured measures of cardiovascular health. Leveraging this algorithm allows us to unlock unstructured clinical data at scale, to combine with more traditional EHR variables for a more robust analysis.

The Dandelion dataset also allows for up-to-date analyses that do not suffer from the same delays as more traditional survey-based or claims-based analyses, allowing us to focus on the long-term efficacy of approved GLP-1 RAs. Further, our treatment effects were directionally consistent with differing modeling approaches and were robust to the specification used.

There are several limitations to note in this study. The most significant limitation was the absence of external pharmacy dispense data to confirm that medication orders had been filled over the course of the study, which would provide a clearer indication of adherence to GLP-1 RA use in the Positive cohort. While this study relies on medication orders to assess which treatment was prescribed by a physician, we cannot speak to the downstream behavior at the patient level. Since it is difficult to confirm adherence without accompanying dispense data for these patients, we anticipate these data to become a part of the Dandelion dataset in the near future.

Another limitation to note is the missing outcomes data at consistent follow-up intervals for all patients. While an RCT has consistent monitoring and more complete follow-up data, real world outcomes data are less consistent and sparse in comparison. We did not find evidence that missingness was associated with any particular patient type or profile, but to the extent that there may be a pattern to the missingness in the availability of covariates or outcomes data, this may impact the magnitude of our findings.

Finally, residual confounding may exist, even though the Positive and Negative cohorts were drawn from the same patient population in the same geographic area during roughly the same time period. Unobserved differences in motivation for obtaining an ECG, initiating GLP-1 RAs, cost of medications given differences in insurance coverage, and other lifestyle factors might play a role. However, given the robustness of our findings, the residual confounding would need to be quite strong to nullify our results, especially for risk attenuation. To the extent that patients who are administered ECGs are receiving them for differing reasons, they are likely similar reasons between the Negative and Positive cohorts, and baseline adjustment for this variation should be effective in mitigating their effect.

### Real-World Impact

Despite these limitations, our results are encouraging as they suggest that expanding access to GLP-1 RAs for a broader population of patients who might benefit from treatment could be very impactful, both in terms of health outcomes for at-risk patients but also with respect to the cost burden faced by healthcare systems. According to an analysis from the Yale School of Medicine, approximately 6.2 million U.S. patients might benefit from taking semaglutide under the new guidance provided by the FDA in response to the results from the SELECT trial. [Lu 2024] These individuals do not suffer from diabetes but are obese or overweight and have histories of preexisting severe cardiovascular disease. With an AI-driven approach using multimodal clinical data to test the impact of less restrictive inclusion criteria, our analysis suggests that the number of patients who could benefit from a GLP-1 RA is underestimated and could be up to 7-fold higher, as many patients treated successfully in our study did not fall into diagnostic categories for which these agents are approved. Extrapolating these findings to the US general population suggests that the potential cardioprotective effects of GLP-1 RAs for primary prevention may potentially extend to 44 million patients nationally, if those who met the broader criteria irrespective of cardiovascular disease history were also eligible and had their risk assessed by ECGs.

If this expanded population of patients were to initiate treatment, this could result in potentially over 17,300 fewer myocardial infarctions and 16,700 fewer strokes annually in the US given the degree of MACE risk reduction observed. [Tsao 2023] Based on the average cost of treating myocardial infarctions and strokes, this could in turn lead to an estimated $490 million in savings per year due to preventing MACE episodes amongst overweight and obese patients. [Khan 2021; Mallow 2023]

## CONCLUSIONS

In this study, we explored how pairing multimodal RWD with an AI algorithm can offer a transformative approach to cardiovascular risk assessment by generating predictive outcomes, in this case to detect early, subclinical changes in cardiovascular health and quantify the risk of MACE using a 12-lead ECG output. We validated that this algorithm performs accurately when compared against ground-truth outcomes, and then leveraged the AI-derived outcomes to conduct a study on the potential cardioprotective benefits of GLP-1 RAs. By focusing on early risk prediction rather than waiting for critical events to occur, our model provides meaningful insights into the cardioprotective effects of GLP-1 RAs in a more diverse and broader patient population than previously studied in the clinical trial setting. As a result, this approach of pairing multimodal RWD with validated AI can potentially streamline clinical evaluations and future clinical trials, reduce costs and risk associated with conducting clinical trials, and expedite the identification of patients who may benefit from early therapy with a GLP-1 RA and other interventions that might be evaluated using this method. The resulting shift away from reactive and toward preventive care offers the potential to improve patients’ lives and reduce the societal and economic burden of cardiovascular disease.

## Data Availability

All data used in this study are available to Dandelion Health AI users.

## REFERENCES

1. Andersen A, Lund A, Knop FK, Vilsbøll T. Glucagon-like peptide 1 in health and disease. Nat Rev Endocrinol. 2018 Jul;14(7):390–403. doi: 10.1038/s41574-018-0016-2

2. Chen Z, Liang N, Zhang H, et al. Harnessing the power of clinical decision support systems: challenges and opportunities. Open Heart. 2023 Nov 28;10(2):e002432. doi: 10.1136/openhrt-2023-002432.

3. Chopra H, Annu, Shin DK, et al. Revolutionizing clinical trials: the role of AI in accelerating medical breakthroughs. Int J Surg. 2023 Dec 1;109(12):4211–4220. doi: 10.1097/JS9.0000000000000705

4. Dandelion Health AI. Syosset, NY. https://dandelionhealth.ai/

5. Hernandez-Boussard T, Monda KL, Crespo BC, Riskin D. Real world evidence in cardiovascular medicine: ensuring data validity in electronic health record-based studies. Am Med Inform Assoc. 2019 Nov 1;26(11):1189–1194. doi: 10.1093/jamia/ocz119

6. U.S. Department of Health and Human Services. Standards for Privacy of Individually Identifiable Health Information; Final Rule.“Code of Federal Regulations, 45 CFR 164.514(b). https://www.hhs.gov/hipaa/for-professionals/privacy/laws-regulations/index.html Hols t JJ. The physiology of glucagon-like peptide 1. Physiol Rev. 2007 Oct;87(4):1409-39. doi: 10.1152/physrev.00034.2006

7. Khan SU, Khan MZ, Khan MU, et al. Clinical and economic burden of stroke among young, midlife, and older adults in the United States, 2002-2017. *Mayo Clin Proc Innov Qual Outcomes*. 2021; 5(2):431-441. doi: 10.1016/j.mayocpiqo.2021.01.015

8. Kong HJ. Managing unstructured big data in healthcare system. Healthc Inform Res. 2019 Jan;25(1):1–2. doi: 10.4258/hir.2019.25.1.1

9. Kostis JB, Dobrzynski JM. Limitations of randomized clinical trials. Am J Cardiol. 2020 Aug 15:129:109–115. doi: 10.1016/j.amjcard.2020.05.011

10. Lee CS, Lee AY. How Artificial intelligence Can Transform Randomized Controlled Trials. Transl Vis Sci Technol. 2020 Feb 12;9(2):9. doi: 10.1167/tvst.9.2.9

11. Li P, Varghese JS, Shah MK, et al. Trends in GLP-1RA Use among Individuals with Type 2 Diabetes (T2D) and/or Obesity—Insights from a Nationwide Electronic Health Record System, 2010–2023. Diabetes. 2024; 73 (Suppl 1): 850-P. doi: 10.2337/db24-850-P

12. Li S, Peng Y, Wang X, et al. Cardiovascular events and death after myocardial infarction or ischemic stroke in an older Medicare population. Clin Cardiol. 2019 Mar;42(3):391–399. doi: 10.1002/clc.23160

13. Lincoff AM, Brown-Frandsen K, Colhoun HM, et al. Semaglutide and Cardiovascular Outcomes in Obesity without Diabetes. N Engl J Med. 2023 Dec 14;389(24):2221–2232. doi: 10.1056/NEJMoa2307563

14. Lu Y, Liu Y, Jastreboff AM, et al. Eligibility for cardiovascular risk reduction therapy in the United States based on SELECT trial criteria: insights from the National Health and Nutrition Examination Survey. Circ Cardiovasc Qual Outcomes. 2024; 17(1):e010640. doi: 10.1161/CIRCOUTCOMES.123.010640

15. Mallow PM, Browne F, Shemisa K. The high cost of death after acute myocardial infarctions: results from a national US hospital database. Clinicoecon Outcomes Res. 2023 Jan 31:15:63–68. doi: 10.2147/CEOR.S397220

16. Marso SP, Daniels GH, Brown-Frandsen K, et al. Liraglutide and cardiovascular outcomes in type 2 diabetes. N Engl J Med. 2016; 375(4):311–322. doi: 10.1056/NEJMoa1603827

17. Marso SP, Bain SC, Consoli A, et al. Semaglutide and cardiovascular outcomes in patients with type 2 diabetes. N Engl J Med. 2016; 375(19): 1834–1844. doi: 10.1056/NEJMoa1607141

18. McGuire DK, Marx N, Johansen OE, Inzucchi SE, Rosenstock J, George JT. FDA guidance on antihyperglyacemic therapies for type 2 diabetes: One decade later. Diabetes Obes Metab. 2019 May;21(5):1073–1078. doi: 10.1111/dom.13645

19. Rajkomar A, Oren E, Chen K, et al. Scalable and accurate deep learning with electronic health records. NPJ Digit Med . 2018 May 8:1:18. doi: 10.1038/s41746-018-0029-1

20. Sandoval DA, D’Alessio DA. Physiology of proglucagon peptides: role of glucagon and GLP-1 in health and disease. Physiol Rev. 2015; 95(**2**):513–548. doi: 10.1152/physrev.00013.2014

21. Sattar N, Lee MMY, Kristensen SL, et al. Cardiovascular, mortality, and kidney outcomes with GLP-1 receptor agonists in patients with type 2 diabetes: a systematic review and meta-analysis of randomised trials. Lancet Diabetes Endocrinol. 2021; 9:653–662. doi: 10.1016/S2213-8587(21)00203-5

22. Tsao CW, Aday AW, Almarzooq ZI, et al. Heart disease and stroke statistics-2023 update: a report from the American Heart Association. Circulation. 2023; 147(8):e93-e621. doi: 10.1161/CIR.0000000000001123

23. U.S. Department of Health and Human Services. Standards for Privacy of Individually Identifiable Health Information; Final Rule.” Code of Federal Regulations, 45 CFR 164.514(b). HHS.gov. Accessed September 10, 2024. https://www.hhs.gov/hipaa/for-professionals/privacy/laws-regulations/index.html

24. Wegovy (semaglutide injection 2.5 mg) [package insert]. Plainsboro, NJ: Novo Nordisk; 2024.

25. US Food and Drug Administration. FDA approves first treatment to reduce risk of serious heart problems specifically in adults with obesity or overweight. FDA Website. March 8, 2024. Accessed August 26, 2026. https://www.fda.gov/news-events/press-announcements/fda-approves-first-treatment-reduce-risk-serious-heart-problems-specifically-adults-obesity-or

